# Microbiomes without boundaries: cystic fibrosis ‘pulmotype’ classifications are dependent on algorithm choice and database size, and indicate continuous variation

**DOI:** 10.1101/2025.05.07.25326893

**Authors:** Conan Y. Zhao, Ryan Lowhorn, Haojun Song, Jinyoung Eum, Sam P. Brown

## Abstract

A common response to microbiome sample variation is to use clustering algorithms to reduce complex and variable datasets to a smaller number of ‘types’ (e.g. enterotypes for gut samples, or pulmotypes for lung samples). In light of recent analyses showing distinct clustering solutions to in principle similar datasets, we examine the extent to which clustering solutions are dependent on researcher choices of algorithm and dataset, using cystic fibrosis (CF) sputum microbiome data as a model system. Following a structured literature review, we identified 36 CF microbiome studies with publicly available samples and metadata. From these studies we curated a dataset of 4026 sputum microbiome samples across 1184 people with CF (pwCF), complete with matched individual metadata, using a standardized bio-informatic platform. Applying multiple clustering algorithms (DMM, k-means, PAM) to cross-sectional data we find that the optimal clustering varies with both algorithm choice and database size, with generally weak separation among clusters in any classification. Our longitudinal data analyses highlight substantial persistence of cluster types in time, with transitions most common among clusters that are structurally similar, reflecting an underlying continuous landscape of microbiome variation. While transitions among similar clusters are common (e.g. along gradients of *Pseudomonas aeruginosa* relative abundance), transitions are generally bi-directional, with no clear pathogen-dominated ‘end point’ states. Using samples from 482 pwCF with available lung function data, we find that taxon-based models outperform cluster-based statistical models in predicting clinical lung function data. Together our results highlight that clustering methods can impose arbitrary boundaries on an underlying continuum of microbiome variation.

**Importance:** Classifying microbiome samples into discrete “types” is a widely used strategy for simplifying complex microbial community data and linking community structure to clinical outcomes. Here we evaluate the utility of cluster-based microbiome typing schemes, using cystic fibrosis (CF) sputum samples as a model system. We conduct a comprehensive re-analysis of over 4000 sputum samples from more than 1000 people with CF. We show that pulmotype classification outcomes are highly sensitive to the choice of clustering algorithm and dataset size, and that clustering can impose artificial boundaries on a continuous landscape of microbial variation. Our findings urge caution over the use of discrete microbiome classifications and emphasize the value of taxon-based models in capturing the ecology and clinical relevance of complex microbial communities.

## Introduction

Microbiome samples from natural (including human) sources are often strikingly variable, even when sampled from seemingly similar locations. For example, fecal microbiome samples can vary substantially across people, even among healthy individuals(Arumugam et al., 2011). One approach to simplify this variation is to cluster individual samples into a smaller number of representative modal ‘types’. This clustering approach has been extensively used in the gut microbiome literature to define ‘enterotypes’(Arumugam et al., 2011, Knights et al., 2014, Costea et al., 2018), and more recently has been applied to lung microbiomes under the term ‘pulmotypes’, notably in the context of people with cystic fibrosis(Widder et al., 2022, Hampton et al., 2021).

Cystic fibrosis (CF) is a genetic disease affecting more than 30,000 people in the US and approximately 70,000 people globally. CF is characterized by chronic, polymicrobial infections of the respiratory tract. Culture-independent sequencing of sputum samples from people with CF (pwCF) have identified thousands of taxa including canonical CF pathogens such as *Pseudomonas aeruginosa* and taxa from commensal oral and nasopharyngeal microbiomes(Caverly et al., 2015, Widder et al., 2022, Hampton et al., 2021). While commonly observed in sputum samples, the functional role of these commensal taxa remains a topic of ongoing debate. Many observational CF studies report a positive association between commensal abundance and lung function(Coburn et al., 2015, Cuthbertson et al., 2016, Widder et al., 2022, Zhao et al., 2012, Zhao et al., 2020) consistent with a beneficial role of commensals. However, experimental *in vitro* microbiome studies show mixed results, with commensals either facilitating or inhibiting pathogen growth, depending on environmental context or species involved(Flynn et al., 2016, Varga et al., 2022). Additionally, the potential for salivary and oral microbiome contamination during sputum expectoration adds a layer of complexity to pulmonary microbiome analyses(Goddard et al., 2012, Jorth et al., 2019), although for patients with more advanced disease this contamination likely has a limited impact(Lu et al., 2020).

To address the substantial variation across CF microbiomes, two studies have applied clustering techniques to categorize these microbiomes into distinct “pulmotypes”. Hampton et al. (Hampton et al., 2021) used k-means clustering on sputum samples from 167 pwCF, identifying five distinct community types which included three dominated by *Pseudomonas*, one dominated by oral taxa, and one mixed-pathogens pulmotype that pooled together diverse cases of established CF pathogens (*Achromobacter*, *Burkholderia*, *Haemophilus*). Widder et al. (Widder et al., 2022) employed a Dirichlet Multinomial Mixture model (DMM) to classify 818 sputum samples across a cohort of 109 pwCF into eight pulmotypes, including separate clusters dominated by *Burkholderia* and *Staphylococcus* along with multiple *Pseudomonas* dominant clusters. Such classification approaches aim to reduce high-dimensional data into more manageable groupings, allowing simpler health outcome comparisons and characterizations of disease progression. Furthermore, defining ‘pulmotypes’ provides a rationale for the development of *in vitro* synthetic microbiomes that potentially represent clinically relevant microbiome structures(Varga et al., 2022, Jean-Pierre et al., 2023).

In light of contrasting pulmotyping methods in the literature(Widder et al., 2022, Hampton et al., 2021), critical questions arise on the robustness of the classifications, and in particular their dependence on the choice of data and algorithm. Individual studies often use relatively small sample sets and varying methodologies, which complicates cross-study comparisons. To address these challenges, we aimed to compile all publicly available CF sputum microbiome data to evaluate whether existing clustering approaches yield consistent classifications across differing choices of algorithm and dataset size.

In this study, we integrate publicly available 16S microbiome data from over 4,000 sputum samples from more than 1,000 pwCF. Beginning with raw reads from NCBI, we applied a standardized bioinformatics pipeline to mitigate methodological discrepancies in data processing. Using this comprehensive dataset, we show that the optimal clustering scheme is highly sensitive to choices of clustering algorithm, with optimal cluster number varying from 2 to 12 on the same dataset. We further show that for some algorithms, the optimal scheme is also strongly sensitive to database size. Using both cross-sectional and longitudinal analyses, we highlight that various cluster partitions generate arbitrary boundaries on a continuously varying landscape of variation. We review the utility of cluster algorithms in light of these results, highlighting the limitations of discrete cluster assumptions given a continuously varying landscape of CF microbiome samples.

## Results

### A Standardized CF Microbiome Database

Using a structured search of NCBI SRA for CF lung microbiome 16S amplicon reads (see methods), we curated a database of 4026 sputum compositions across 1184 individuals with CF from 36 published studies (median 53 samples per study; 1-163 samples per individual pwCF), Table 1. Our database uses a common read processing pipeline (see methods) and represents 26 CF centers across 14 countries (Figure 1A). Given the variation in both inclusion criteria and methodological choices within each component study, we hypothesized that methodological and regional differences will result in compositional differences among studies. Consistent with this hypothesis, we see significant differences in composition in multiple pairwise study comparisons (Figure 1B, Analysis of Similarity ANOSIM R *p* < 0.05 Bonferroni corrected), yet effect sizes rarely approached our threshold for a substantial effect (ANOSIM R > 0.4(Roberts et al., 2008)).

**Figure 1.**
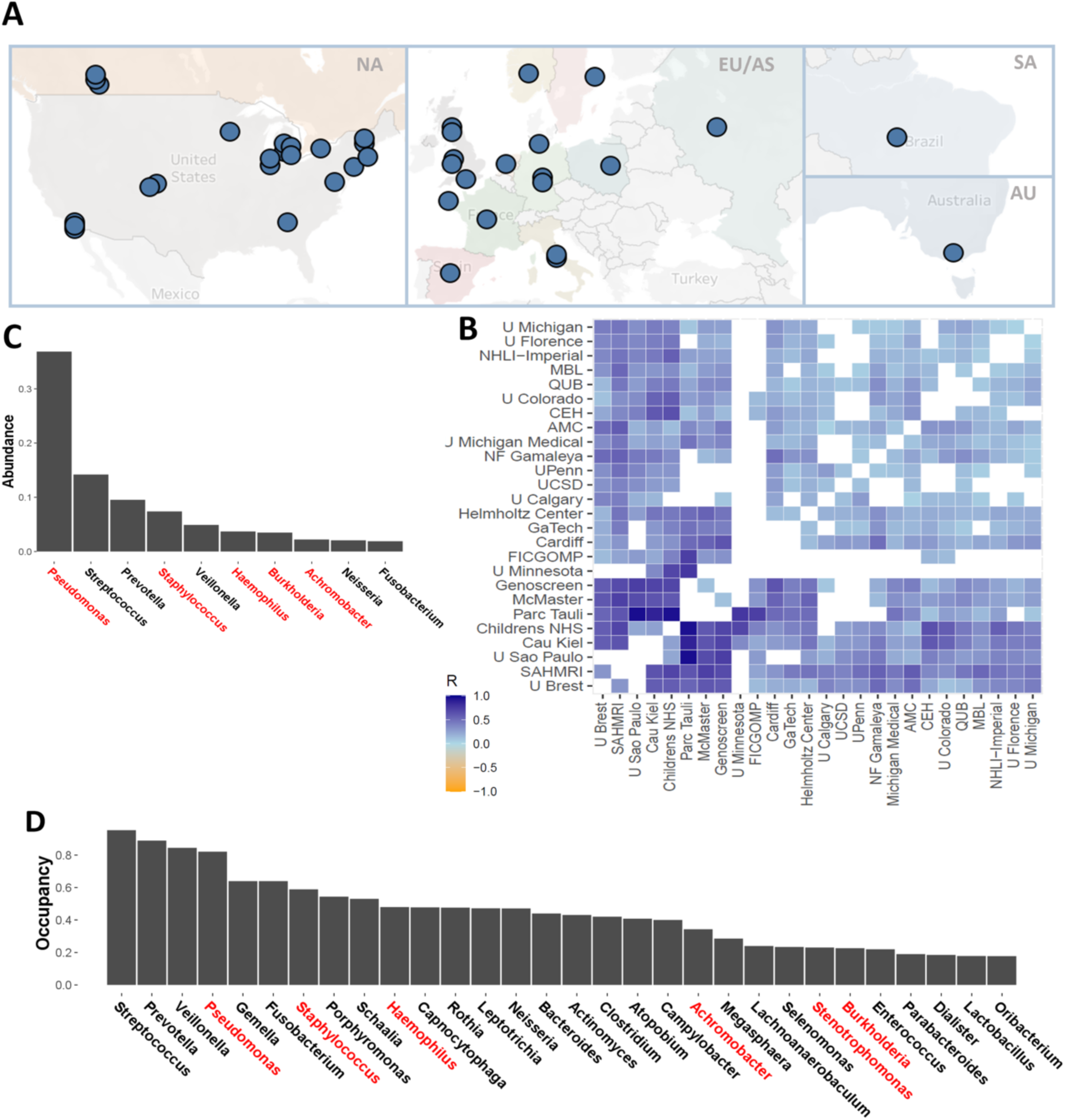
A multi-study CF database spanning 1184 pwCF and 4171 sputum samples. (**A**) We analyzed 36 studies across 26 CF centers from 14 countries worldwide. Three studies (PRJEB30646, PRJEB38277, and PRJNA207555) are split across more than one study site. (**B)** Pairwise ANOSIM R statistic for studies grouped by CF center that uploaded the study. White spaces denote pairs that were not significantly different (p > 0.05, Bonferroni corrected). (**C**) Mean relative abundance by genera. (**D**) Mean prevalence by genera. Canonical CF pathogens (*Pseudomonas, Staphylococcus, Burkholderia, Stenotrophomonas, Achromobacter,* and *Haemophilus*) are highlighted in red.

**Table 1.**
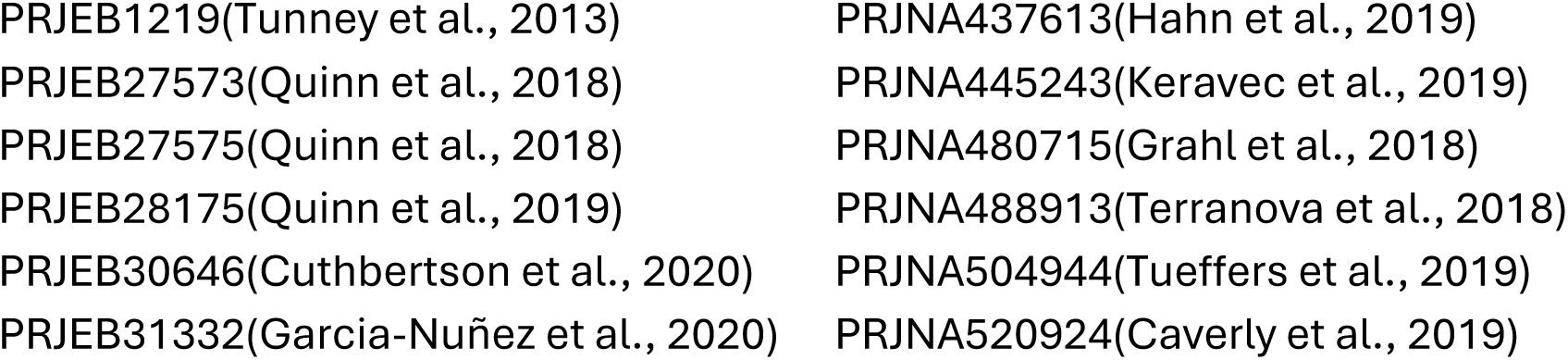
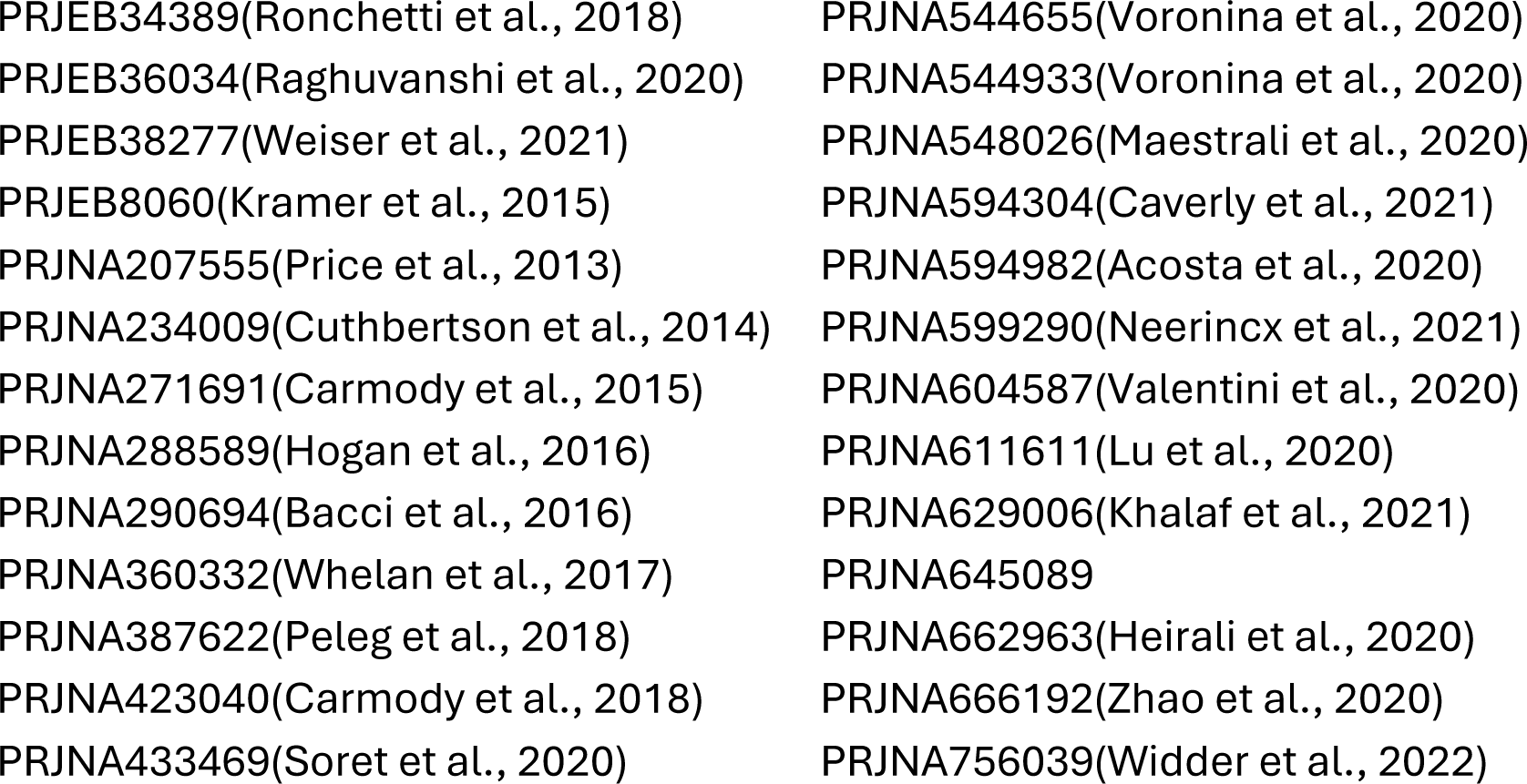
NCBI Bioproject numbers and linked references, where available. Data available via https://www.ncbi.nlm.nih.gov/bioproject.

Post-rarefaction, 95% of reads came from 19 taxa. *Pseudomonas* was the most abundant taxa detected, followed by *Streptococcus*, *Prevotella*, and *Staphylococcus* (Figure 1C). *Streptococcus* was the most prevalent taxa, occurring in 95.3% of all samples, followed by *Prevotella*, *Veillonella*, and *Pseudomonas* (Figure 1D).

### 1184 CF microbiomes partition into 12 ‘pulmotypes’ via DMM

To avoid over-representation by longitudinally tracked pwCF, we begin with a purely cross-sectional approach, taking only the first sample from each of the 1184 pwCF in our complete dataset. In brief, we filtered taxa by prevalence across studies (74 genera retained) and then used the specific DMM clustering algorithm used previously(Widder et al., 2022) to generate an 8 pulmotype schema (see methods for detailed processing and analysis steps).

Comparing the performance of one to 25 clusters, we find that CF lung microbiomes are best partitioned by DMM into twelve pulmotypes (Figure 2A-C), in contrast to prior studies identifying 5(Hampton et al., 2021) or 8(Widder et al., 2022) pulmotypes. The optimal 12 pulmotype partitioning is detailed in Figure 2C (ordered by abundance), showing the relative abundances of the top-10 genera for each pulmotype. The count distribution across the 12 pulmotypes is shown in Figure 2B, with counts ranging from 16 to 214 samples per pulmotype.

**Figure 2.**
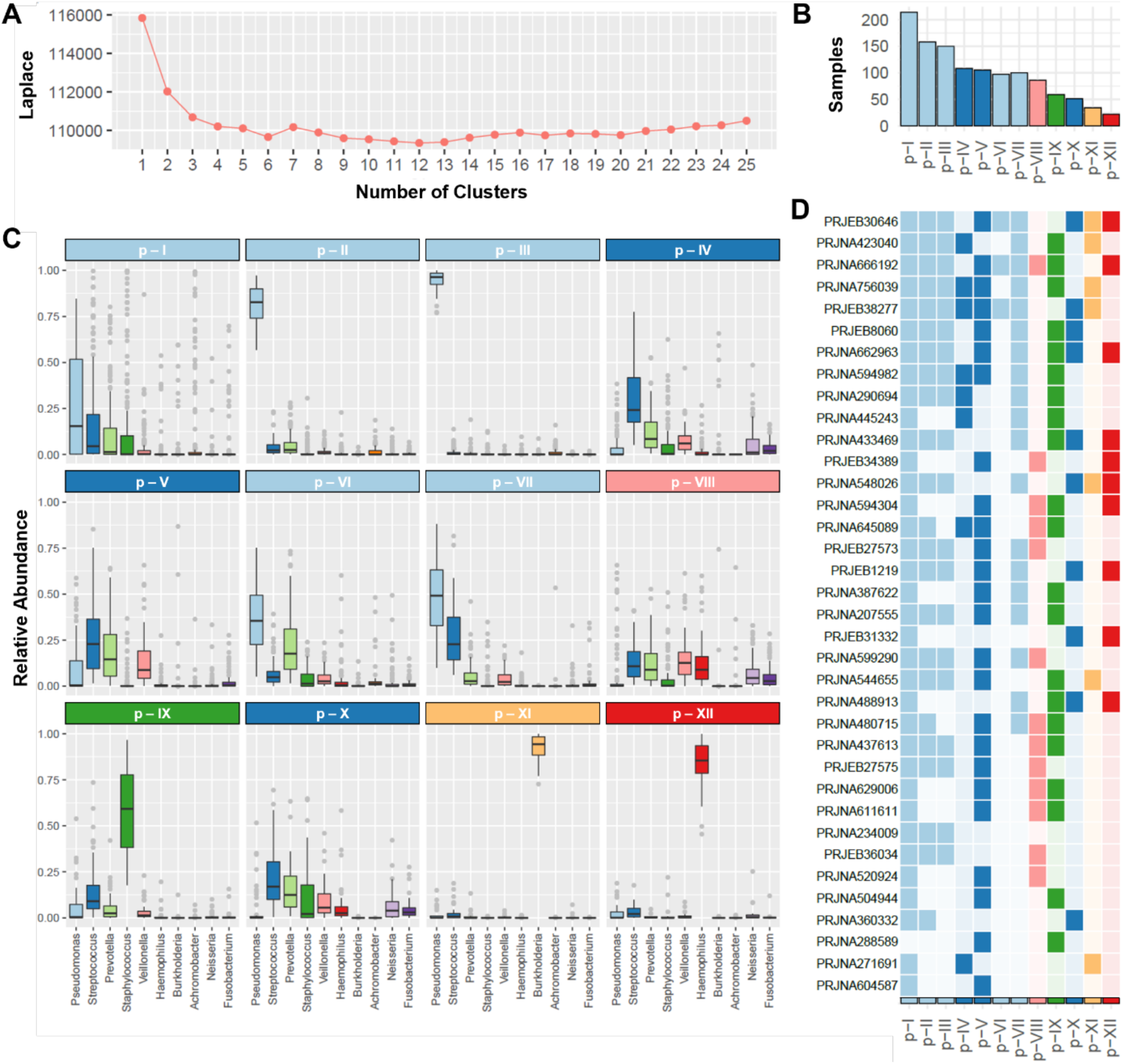
1184 cystic fibrosis sputum microbiomes separate into 12 pulmotypes, via DMM. Clusters (k=1…25) were calculated using DMMs on individual snapshot data (N=1184) of the 74 most prevalent taxa, rarefied to 2000 sequences each. (**A**) Using the Laplace approximation of the negative log model evidence, we find a minimum at k=12 clusters (pulmotypes). (**B**) Overall frequency of each pulmotype across the initial 1184 samples. (**C**) Boxplots of the top 10 taxa grouped by 12 pulmotypes. (**D**) Pulmotypes represented in each study are shaded in. Studies are ordered by number of samples included.

We further organize our 12 clusters into 3 broad groups: **Pseudomonas-dominant (PA), oral commensal dominant (OC),** and **other pathogen dominant (OP).** The majority (60.7%) of the 1,184 samples are categorized as one of five PA-dominant pulmotypes (p-I, p-II, p-III, p-VI, and p-VII). The most common pulmotype is p-I (*n* = 214), which has moderate *Pseudomonas* relative abundance. Pulmotype p-III is the most homogeneous (DMM θ = 40.38; higher θ is more homogeneous) and typically exhibits over 90% *Pseudomonas*. By contrast, p-I is the least homogeneous (θ = 2.01) and contains a wide range of *Pseudomonas* levels. The PA pulmotypes also vary in overall *Pseudomonas* relative abundance (from lowest to highest: p-I, p-VI, p-VII, p-II, p-III). This pattern is consistent with a continuous gradation of *Pseudomonas* across samples arbitrarily clustered into distinct pulmotypes.

The four OC pulmotypes (p-IV, p-V, p-VIII, p-X) comprise 350 (29.5%) samples and are dominated by *Streptococcus* or *Veillonella*. Pathogen levels in the OC pulmotypes are generally low. Meanwhile, the three OP pulmotypes (p-IX, p-XI, and p-XII) total 115 samples (9.7%) and are dominated by non-*Pseudomonas* pathogens— namely *Staphylococcus* (p-IX), *Burkholderia* (p-XI), and *Haemophilus* (p-XII). Notably, we do not find a single OP pulmotype comprising *Stenotrophomonas*-dominated cases, despite loads exceeding 90% in some samples. Instead, *Stenotrophomonas*-heavy samples are absorbed into p-I or p-III (both PA), or one of the OP clusters, rather than forming a separate pulmotype. A risk of our multi-study approach is that clustering will simply separate pwCF by study.

To begin to examine the validity and distinctness of the 12 pulmotypes, we first analyze their representation across included studies. In contrast to a major study effect on classification, we find that every pulmotype is represented in at least six studies, although no study contained all twelve pulmotypes (Figure 2D). The top four studies by number of pulmotypes represented (in decreasing order by total number: PRJNA423040, PRJNA756039, PRJEB30646, and PRJNA666192) were larger cross-sectional studies (pwCF > 57), including two that were used in prior pulmotyping analyses(Widder et al., 2022, Hampton et al., 2021).

### Pulmotype classification is sensitive to database size and algorithm choice

To summarize our initial analyses, the DMM method applied to 1184 pwCF produces 12 clusters that are broadly represented across individual studies (Figure 2). While the DMM pipeline supports a 12 cluster solution, we can see from Figure 2A that other clustering solutions (e.g. 11 and 13) are almost equally well supported. To assess the robustness of the 12 cluster solution, we next experiment by applying the same DMM method to repeated sub-samples of our dataset to establish confidence intervals on cluster number, and also to assess any dependency of cluster number on database size.

Figure 3 illustrates that when we repeat the same DMM clustering analysis on sub-samples of our 1184 pwCF dataset, we see consistency across replicates in cluster number for a given sample size, but also systematic variation as sample size changes (Figure 3). This strong dependency on sample size provides a simple rationale for why our clustering scheme (Figure 2) has more clusters than an earlier study applying the same algorithm to a smaller dataset(Widder et al., 2022), or indeed different algorithms to smaller datasets (Hampton et al., 2021).

**Figure 3.**
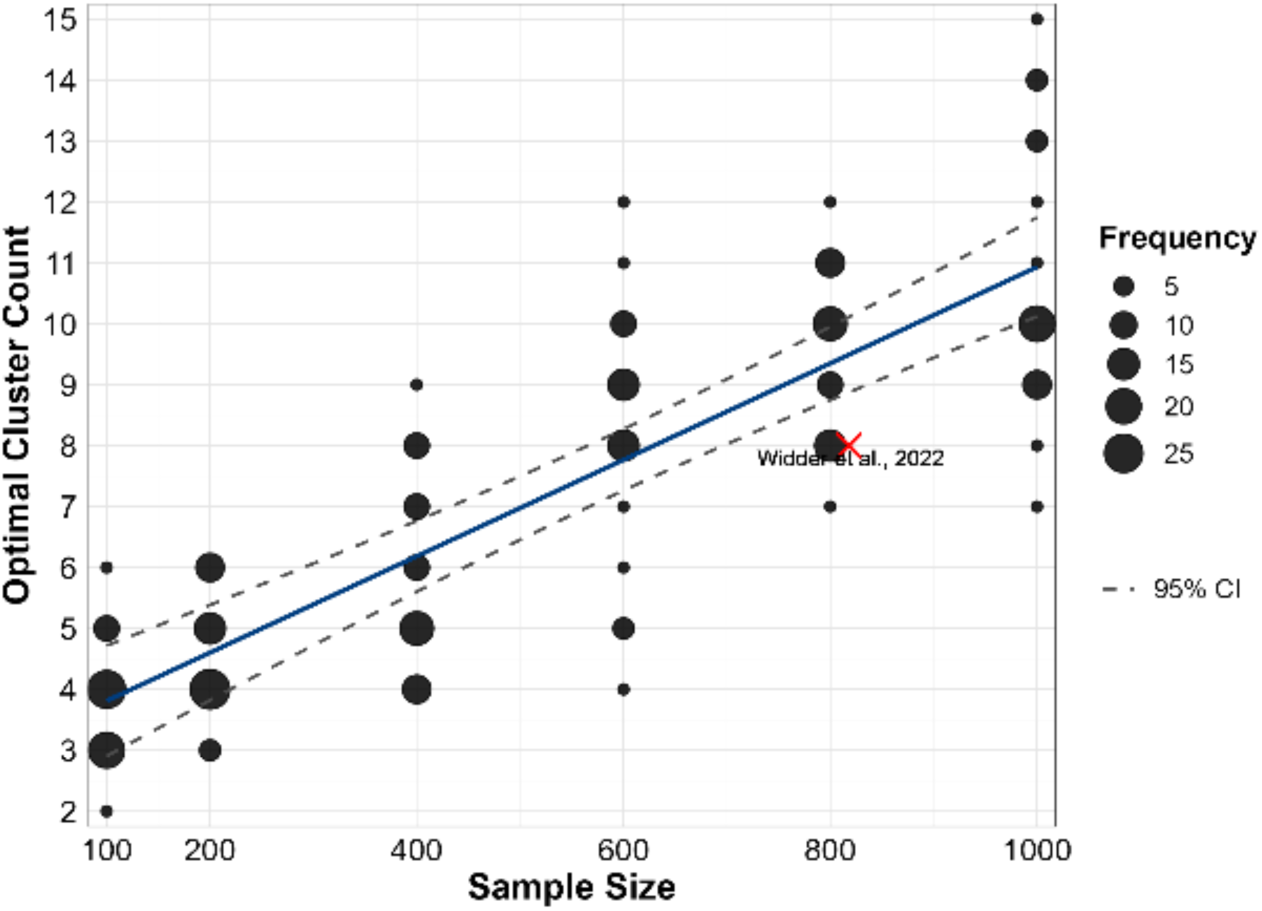
DMM clustering results are dependent on database size. The 1184 cross-sectional database was sub-sampled (without replacement) 60 times, for sample sizes ranging from 100 to 1000 pwCF. Points illustrate the optimal cluster count (analogous to the 12-cluster count in Figure 2), with point size illustrating the number of samples with the same cluster count (see inset). The blue line is a linear regression of mean cluster count on sample size, surrounded by 95% confidence interval (dotted lines). The red cross represents the DMM analysis by Widder et al. (Widder et al., 2022)(DMM clustering applied to 818 longitudinal samples from 109 pwCF).

To assess the potential role of algorithm choice, we next use our 1184 pwCF dataset to contrast DMM with other commonly-used clustering algorithms (k-means, and partitioning around medoids (PAM)). In Figure 4, we show that algorithm choice has a substantial effect on clustering outcome. For the same 1184 pwCF dataset, the resulting number of clusters varies from 2 (PAM) to 12 (DMM). Given the variability in clustering outcomes from a single dataset, an objective evaluation of algorithm performance is essential. We assess within-cluster cohesion and between-cluster separation using the Average Silhouette Width (ASW) metric (Figure 4). Higher ASW scores for PAM indicate a more robust clustering scheme.

**Figure 4.**
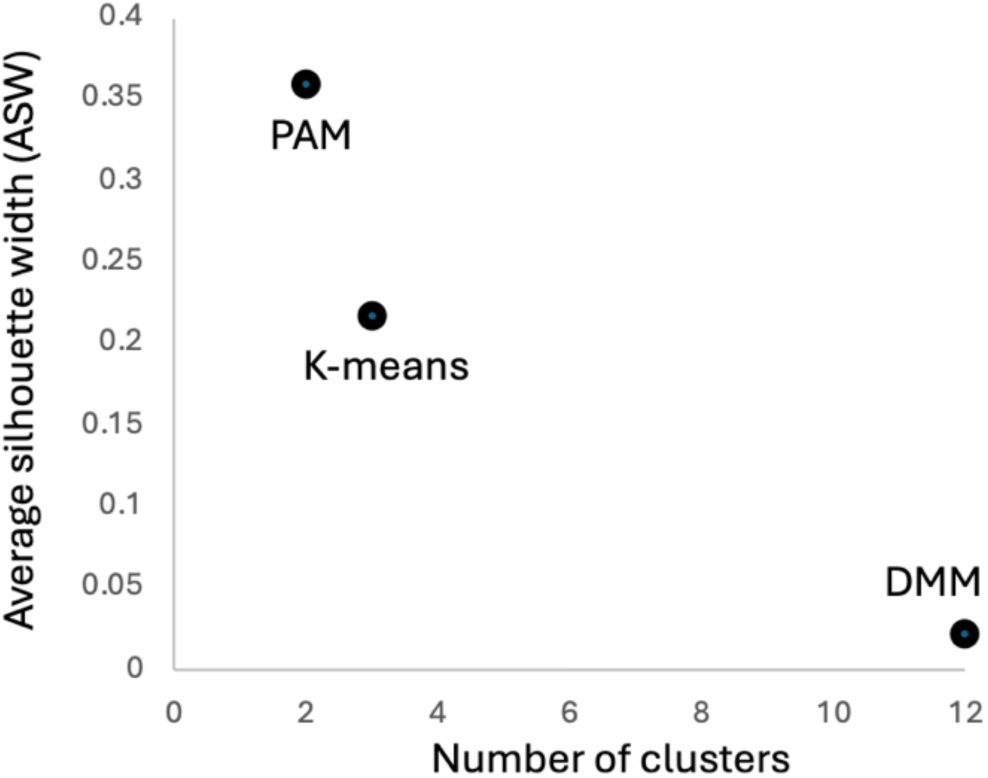
cluster number and quality is dependent on algorithm choice. The complete 1184 cross-sectional dataset was used to fit DMM, k-means and PAM (partitioning around medoids) clustering models. The x-axis indicates the number of clusters identified, while the y-axis indicates the Average Silhouette Width (ASW) score, a metric evaluating cluster quality, with a range from −1 to 1; higher scores denote stronger clustering (greater within-cluster homogeneity and between-cluster separation).

Our analysis in Figure 4 demonstrates that the PAM two-cluster scheme achieves higher clustering quality, as indicated by higher ASW scores. Building on this, we next evaluate the sensitivity of PAM clustering to database size, given our earlier finding that DMM clustering is size-dependent (Figure 3). As shown in Figure 5, unlike DMM clustering, PAM clustering remains highly robust across database sizes, consistently identifying the same two-cluster structure from 200 to 1000 pwCF. Notably, the composition of these clusters remains highly conserved, further supporting the robustness of this classification.

**Figure 5.**
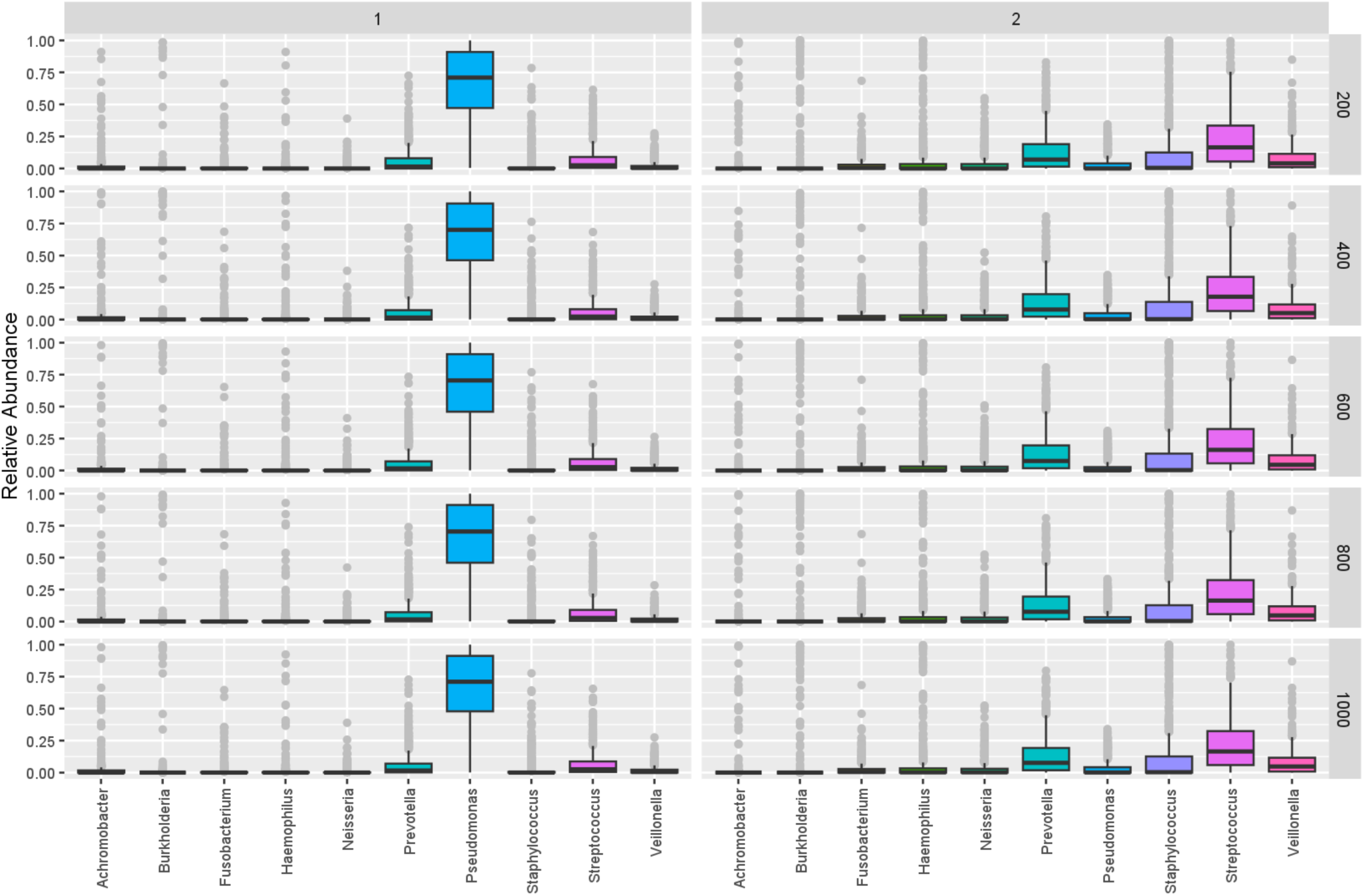
PAM clustering produces a repeatable 2 cluster schema, across database sizes. PAM clustering was repeated on single subsamples of our data, producing analyses on datasets ranging from 200 (top) to 1000 pwCF. For each PAM analysis, the optimal clustering partitioned the data into two clusters, with compositions detailed in the left panel (cluster 1) and the right panel (cluster 2). Each panel shows the relative abundance for the top 10 most abundant genera. The boxplots depict the variation in abundance of each genus within the identified clusters, with outliers indicated by grey dots.

Our analyses in Figures 4-5 indicate that a two-cluster model identified by PAM provides a robust classification scheme based on available cross-sectional data. Compositionally, this two-cluster model separates PA-dominant microbiomes (cluster 1, Figure 5) from all others (cluster 2). This distinction is consistent with the clinical weight placed on PA infections (Emerson et al., 2002) and is partially consistent with prior models differentiating ‘attack’ and ‘climax’ microbiome structures in pwCF, where PA has been associated with the climax community(Quinn et al., 2016, Conrad et al., 2013).

### Clusters versus continua

Clustering algorithms always generate clustering solutions, regardless of whether the underlying data naturally forms discrete groups. Our ASW metric provides some insights into this issue, offering relative support for PAM clustering. Yet even in this most favorable case, the measured ASW is still relatively weak (below a suggested threshold of 0.5(Knights et al., 2014)), suggesting model misspecification. Since our PAM two-cluster model separates by PA abundance (Figure 5), we next assess the extent to which PA abundance forms a continuous (graded) or discrete (step-like) landscape across all 1184 initial timepoint samples (Figure 6).

**Figure 6.**
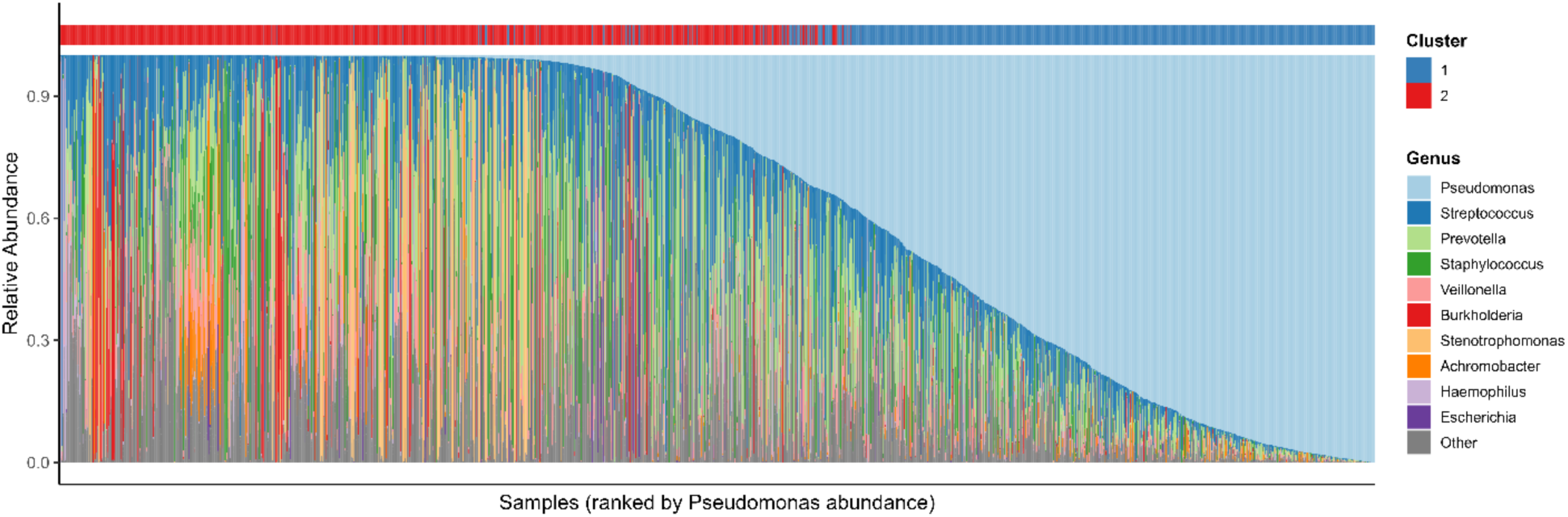
The PAM two cluster classification does not reflect a sharp boundary in *P. aeruginosa* abundance. Microbial community composition across samples ranked by *Pseudomonas* (PA) abundance. Each vertical bar represents a single sample, and the stacked colors within each bar correspond to the relative abundances of the top 10 most abundant bacterial genera, with all other genera combined into the “Other” category. Samples are ordered from left to right by increasing PA abundance (pale blue). A colored strip above the composition bars indicates the cluster assignment derived from partitioning around medoids (PAM) clustering.

While we again see that clusters 1 and 2 are largely separable by PA relative abundance (Figures 5, 6), figure 6 shows that there is no sharp threshold in the abundance of PA across all samples, suggesting a continuous underlying landscape of variation.

### Transition patterns in longitudinal data reveal a continuous landscape connecting DMM clusters

Figures 2-6 show our analyses of initial timepoint data for 1184pwCF, revealing limitations for all clustering models. Broadly, our analysis points to a choice between a high-resolution twelve-cluster DMM model (but with low ASW scores and high sensitivity to dataset size, Figures 2-4), or a simple two-cluster PAM model (but with only marginal ASW improvement, Figures 4,5). In figure 6 we critique the underlying ‘discrete space’ pulmotyping assumption, by illustrating a clear compositional continuity across the major two-cluster border in the PAM model.

We now move on to our longitudinal sample dataset, to further evaluate a continuous space interpretation, where clusters partition an underlying continuous landscape of microbiome variation. Under this interpretation, we hypothesize that longitudinal transitions in pulmotype cluster reflect underlying distances across a continuous microbiome landscape. To test this hypothesis, we focus on our twelve-pulmotype DMM partitioning (Figure 2), analyzing transitions within and between clusters, and cluster network properties. We use the 12-pulmotype DMM partition (Figure 2) as it provides a larger number of between-cluster transitions, also varying in cluster distances.

Using the prior 12-component DMM, which was fit to cross-sectional data alone, we classify the remaining 2,842 longitudinal samples (across 427 pwCF). Note that while we can infer ordinality (i.e. “which sample came first, second, etc…”) across all subjects with longitudinal samples, we do not always have temporal information (i.e. “how many days passed between samples”). Among all pairs of consecutive samples (2,842 total), 1,686 (59.3%) show no change in pulmotype. Across all individual pwCF in our longitudinal dataset, we find that 125 pwCF (29.3%) do not transition to a new pulmotype during their surveillance period, and 197 pwCF (46.1%) do not change their pulmotype group (i.e. remaining with the broad classification of PA, OC, or OP dominated pulmotypes). Taken together, this indicates stability across pulmotype classification.

We next analyze the characteristics of pulmotype transitions between consecutive samples (Figure 7). For each of the 144 possible ordered pairs of pulmotypes we report the number of individuals in which we find that given transition across two consecutive samples (Figure 7A). We hypothesized that transitions in pulmotype state would result from either misclassification (samples drifting along pulmotype boundaries) or progressive (directional) community shifts. If misclassification is a substantial issue, we would expect to see a negative association between assignment confidence (the likelihood of observing a given sample in its assigned DMM cluster) and the likelihood of a shift in pulmotype in the subsequent sample. In contrast, we find that within-pulmotype and between-pulmotype transitions do not differ in sample-pulmotype assignment confidence (p=0.12, Wilcoxon W=20545, Figure 7B), suggesting that pulmotype transitions are not substantially due to misclassification.

**Figure 7.**
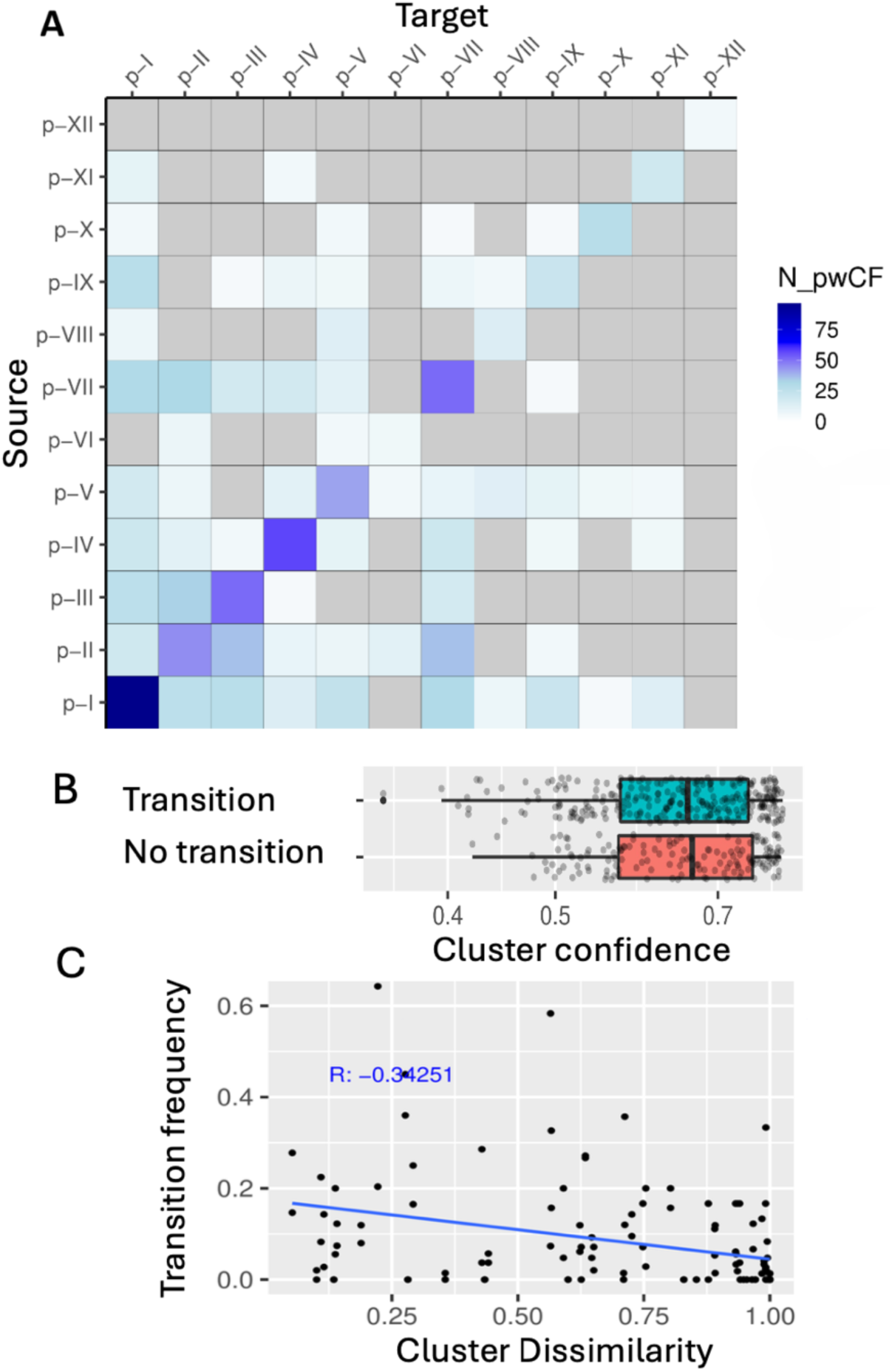
DMM pulmotype assignments are persistent in time, with greater transitions among more similar pulmotypes. We assess transition frequencies between pairs of consecutive samples for each pwCF. (**A**) Matrix of all transitions between pulmotypes, colored by the number of individuals a given transition was observed in. Following Widder et al.(Widder et al., 2022), all transitions with fewer than 3 observations were removed. (**B**) The pulmotype assignment confidence (the likelihood of observing a given sample in its assigned DMM cluster) is not different in between-pulmotype transitions compared to within-pulmotype transitions. (**C**) The transition frequency between two pulmotypes decreases with mean dissimilarity (ANOSIM R).

We next assess whether our classification supports a proximity model, where transitions are more common among pulmotypes that are more structurally similar (and thus have a smaller between-pulmotype distance). Consistent with this model we find that pairs of pulmotypes with lower Bray-Curtis dissimilarity experience more frequent transitions (R=-0.34, Figure 7C).

In light of Figure 7, we conclude that transitions among DMM clusters are in part a function of cluster distance, so that ‘nearby’ clusters (lower Bray Curtis dissimilarity) are more likely to experience transitions (Figure 7D). We build on this interpretation by using cluster transition data (Figure 7A) to build a cluster transition network, where cluster-pairs with more frequent transitions are linked by a network edge (Figure 8). We threshold our network edges based on pulmotype assignment confidence, only including transitions where the product of source and target confidences exceeded 80%. This removes 819 transitions (28.8% of total transitions). Edges in the network are scaled by the number of pwCF for which a given transition was observed. Node sizes are scaled by frequency across the initial DMM training set (Figure 2B).

**Figure 8.**
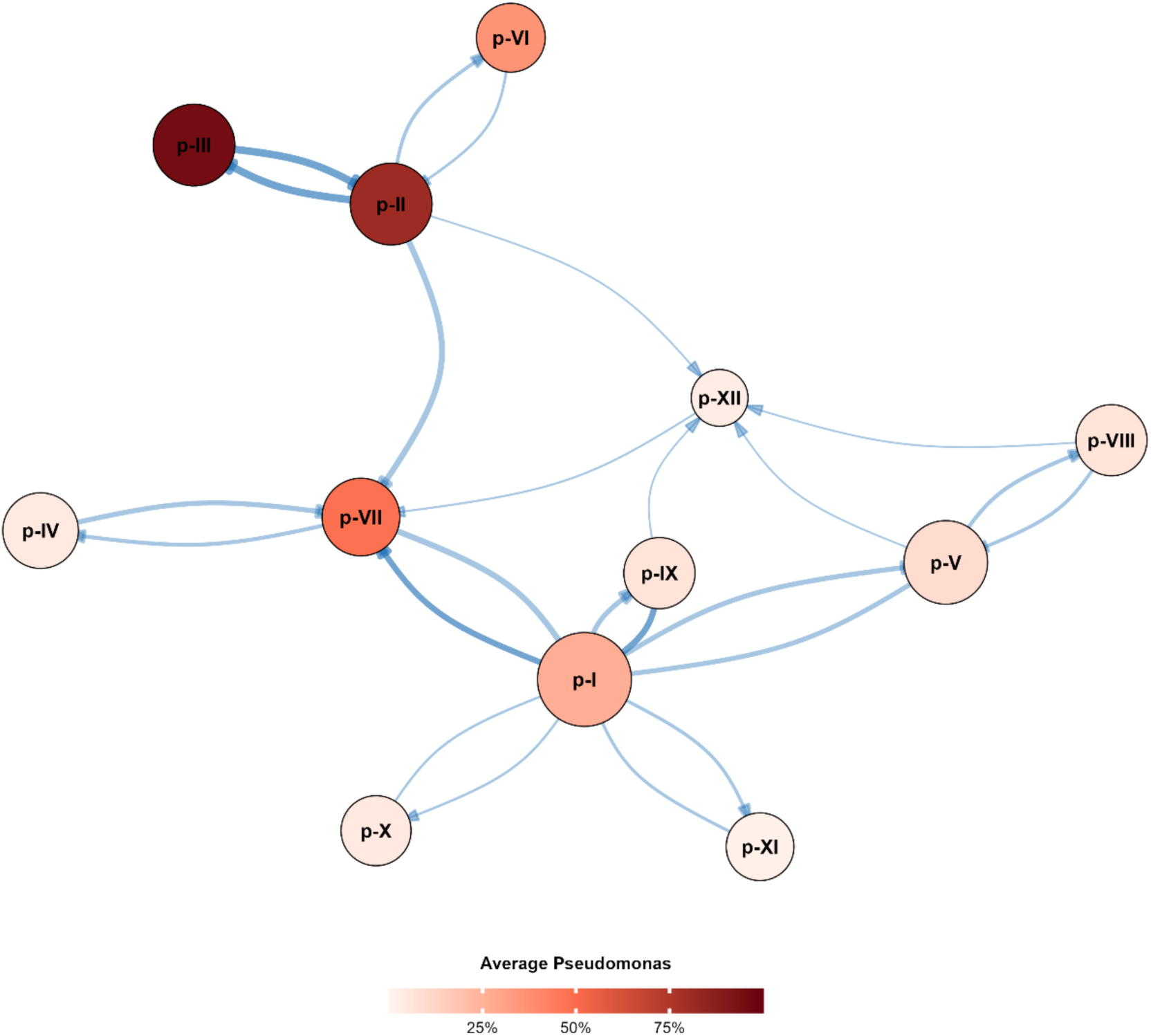
DMM pulmotype transitions form a structured bi-directional network, reflecting gradients in *P. aeruginosa* relative abundance. The network diagram shows each of the 12 DMM clusters as a node, with edges indicating observed transitions between clusters (thresholded by product of source and target assignment confidence > 80%). The node size reflects how many total samples were assigned to that cluster (larger nodes = more samples), and the node color is a gradient representing the average abundance of *Pseudomonas aeruginosa* in that cluster (darker indicates higher *Pseudomonas* levels, see heatmap inset). The edges are drawn with widths proportional to the transition frequency between clusters, and arrowheads denote the direction of the transition.

Our network visualization in Figure 8 reveals a backbone of ordered transitions in PA-dominant pulmotypes along a gradient of *P. aeruginosa* relative abundance, consistent with our earlier illustration of a continuous landscape of *P. aeruginosa* abundance (Figure 6). Figure 8 highlights that movement along this continuous gradient tends to be gradual, so that transition events tend to favor small step changes in *P. aeruginosa* abundance. Across all transitions in Figures 7 and 8, we find substantial bi-directionality (forward and reverse transition rates), indicating a general absence of progressive, directional change in pulmotypes through time.

Figures 7 and 8 show frequent transitions from pathogen dominated (OP or PA) to oral commensal (OC) clusters, suggesting that pathogen dominated clusters are not always ecological sinks (i.e., pwCF can and do escape from these states).

### Taxon-based models can outperform cluster-based models in explaining variance in clinical outcome data

In a final data analysis step, we turn to the critical question of whether pulmotyping can enhance our ability to explain variation in patient outcomes. Hampton et al. (Hampton et al., 2021) showed that their 5-pulmotype schema explains substantially more variance in clinical lung function data than established predictors of health outcomes (*P. aeruginosa* infection, patient age, microbiome Shannon diversity – alone and in combination). Specifically, they found that their model could explain 24% (adjusted *r*^2^ = 0.24) of the variance in ppFEV1 (percentage predicted Forced Expiratory Volume in 1 second).

Here we follow the same adjusted *r*^2^ approach applied to our ensemble dataset for cases where the initial pwCF observation has publicly available data on ppFEV1 (482 pwCF, Figure 9). In Figure 9A, we evaluate different cluster-based models and find the highest adjusted *r*^2^ (*r*^2^ = 0.11) for a DMM partition into 8 clusters. Looking across cluster-based models, we see that model improvement saturates before the 12-cluster DMM (slightly lower *r*^2^), implying that more fine-scale cluster partitioning doesn’t improve model performance when accounting for the increased number of model parameters. Compared to the Hampton study, our adjusted *r*^2^ values are lower, likely reflecting greater clinical heterogeneities in our global composite dataset.

**Figure 9.**
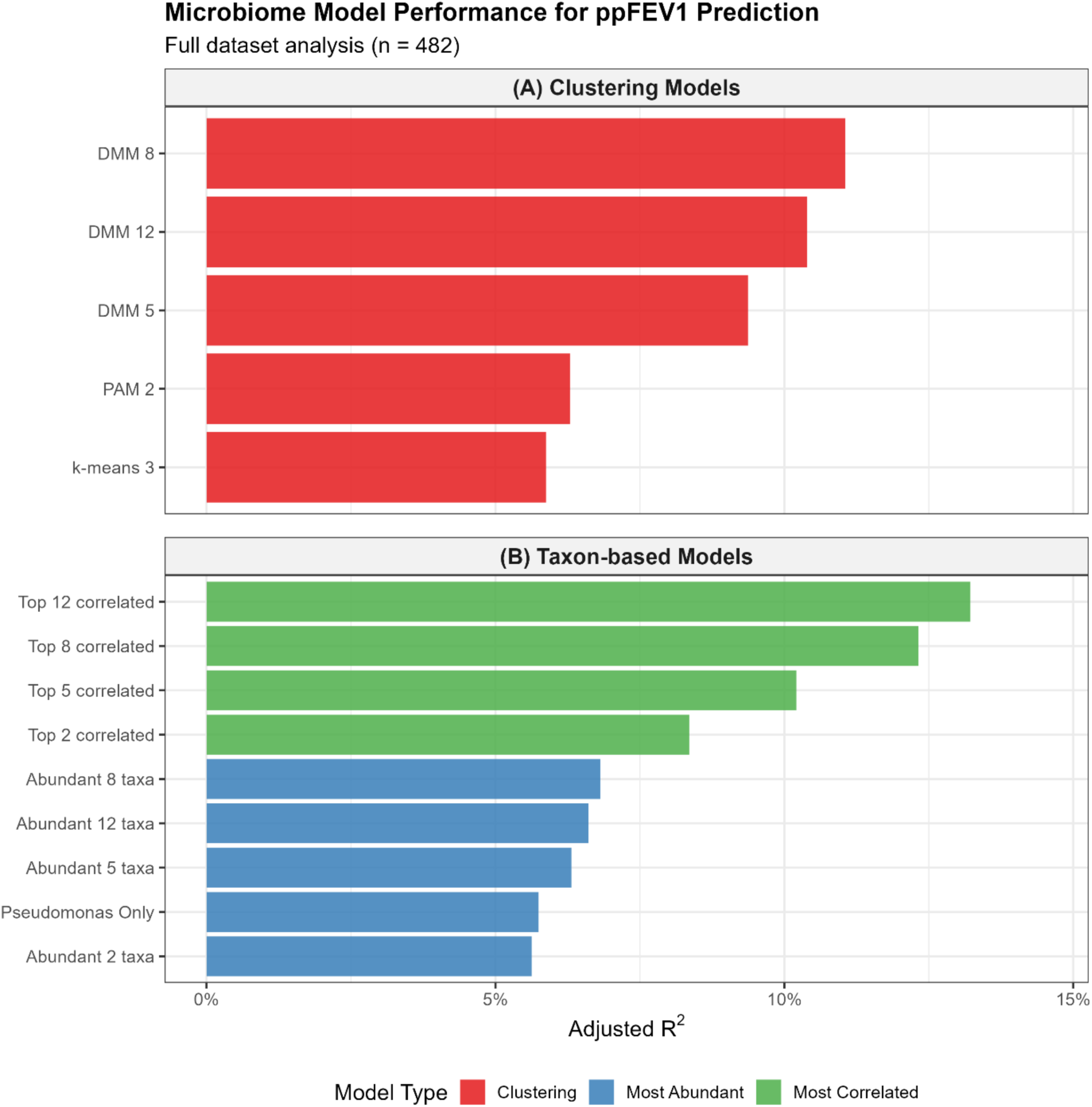
Taxon-based models outperform cluster based models in predicting clinical lung function data. Top panel: cluster-based model performance, assessed by adjusted *r*^2^ (following (Hampton et al., 2021)). The 12-cluster DMM, 2-cluster PAM and 3-cluster K-means are the algorithm-specific optimal clustering solutions (Figures 2,4,5). Bottom panel, taxon-based model performance, using different feature selection approaches (see legend). Specific taxa lists for taxon-based models are detailed in supplementary table S1.

Having established a cluster-based model baseline for our dataset, we next ask in Figure 9B whether equivalently complex taxon-based models can provide better or worse explanatory power. In our first analyses, we select taxa simply by ranked abundance, from *P. aeruginosa* alone (most abundant, Fig 1C), through to top 12 most abundant taxa (blue bars, Figure 9B). Across these models, we see a limited spread of adjusted *r*^2^ values, peaking for the top-8 taxa model (*r*^2^ = 0.07). Previous work has flagged that specific taxon combinations can enhance ppFEV1 predictions when taxa are selected based on their explanatory power(Zhao et al., 2020), so we next used a simple correlation criterion for feature selection, picking the top 2 (or 5, 8 or 12) most correlated taxa with ppFEV1. Using this approach, we see a continuous increase as we add more highly correlated taxa, with adjusted *r*^2^ peaking for the top-12 most correlated taxa (*r*^2^ = 0.13). For each level of model complexity (2, 5, 8 or 12 features), looking across Figures 9A and 9B we see that taxon-based models can outperform cluster-based models.

## Discussion

In this study, we combined publicly available sputum sample microbiome data to identify consistent ecological patterns across over 1000 pwCF. We produce five key findings: (1) cluster partitioning is sensitive to both choice of algorithm and database size (Figures 2-6); (2) longitudinal time-series display clustering stasis (Figure 7); (3) Longitudinal transitions are biased towards similar clusters, e.g. movement along close gradations in *P. aeruginosa* dominance (Figure 7,8); (4) longitudinal cluster transitions show balanced bi-directionality, in contrast to the notion of progressive movement towards ‘end state’ pathogen dominance (Figures 7,8); (5) cluster-based predictive models can be out-performed by equivalently-complex taxon-based models (Figure 9). The database assembled by this study is publicly available via the Zenodo research repository (https://zenodo.org/uploads/15801571) and represents a significant resource to both the CF community and for scientists engaged in the study of human associated microbiomes in the context of disease.

The field of CF has generated a substantial number of microbiome studies. These studies have identified complex microbial communities inhabiting CF airways with high between-person and even within-person variability. In light of the multitude of generally low sample size studies (understandable given the limitations of clinical studies of rare genetic conditions), composite studies are potentially beneficial in identifying broad-scale patterns across numerous cohorts. For example, Li et al. performed a data mining analysis across 18 studies with 718 sputum samples and found that overall, antibiotic treatments for exacerbation management had a large impact on commensal taxa but little impact on CF pathogens(Li et al., 2016).

As reviewed earlier, we are not the first study to apply pulmotyping to CF(Widder et al., 2022, Hampton et al., 2021). Together these variable prior classifications underline our conclusion that clustering solutions are highly sensitive to choice of clustering algorithm and constraints on the amount of available data (Figures 2-5). In the specific case of DMM clustering, our results in Figure 2 compared to Widder et al. (Widder et al., 2022) suggest that more data buys greater resolution of what is fundamentally a continuous landscape of underlying variation (Figure 3).

To conduct a longitudinal analysis we used our 12-cluster DMM partitioning (Figure 2), as it offered a higher cluster number and therefore the potential for more transitions and a more complex transition network (Figures 7,8). Our result that cluster transitions are substantially bi-directional (Figures 7,8) is consistent with (Widder et al., 2022, Widder et al., 2024), and raises the question of what processes drive exits from potentially ‘end stage’ pathogen dominated states (notably DMM clusters II, III (heavy *P. aeruginosa* dominance) and XI (*Burkholderia* dominance). We speculate that the transitions between PA states with higher *Pseudomonas* loads to lower ones correspond to antibiotic treatment, as similar transitions were identified in (Widder et al., 2022, Carmody et al., 2022). These results are consistent with the notion that antibiotic treatment of *Pseudomonas* leads to variable outcomes(Somayaji et al., 2019), potentially either reducing pathogen loads to a more oral commensal-dominant state, or potentially allow for competitive release of other pathogens(Varga et al., 2022, Waldetoft et al., 2022) and therefore promote transitions into distinct pulmotypes. We were ultimately unable to assess this hypothesis given a lack of published, standardized antibiotic information, and we suggest that the field move towards recording and sharing antibiotic prescription data where possible to aid in future studies.

In this paper we have developed and applied a robust pipeline to generate a combined dataset of unprecedented size in CF research, representing over 1000 pwCF and over 4000 samples. Using the dataset, we show that microbiome clustering solutions are dependent on both choice of algorithm and scale of data (Figures 2-6). While specific clustering solutions are variable, we provide evidence that the underlying continuous variation in microbiome structure forms an ordered landscape, characterized by preferential bi-directional transitions to nearby states (Figures 7,8).

## Data Availability

All data analyzed in the present study are open source and available via https://www.ncbi.nlm.nih.gov/bioproject (specific bioproject numbers are detailed in Table 1).

## Acknowledgements

We thank Arlene Stecenko, Joshua Weitz, Peng Qiu, John Li Puma, Thomas Hampton, Bruce Stanton, George O’Toole and members of the Brown lab for helpful feedback on previous drafts. We thank the Cystic Fibrosis Foundation (awards BROWN23I0, BROWN21P0) and the CDC (75D30120C09782) for funding this work.

## Methods

### Dataset Curation

On February 9th, 2022 we searched NCBI-SRA for 16S-sequenced sputum surveys of CF lung microbiomes using the following query:

(((cystic fibrosis OR cf) AND (lung OR respiratory OR sputum OR airway))) AND (amplicon[Strategy] OR other[Strategy]).

This returned 107 potential BioProject numbers. We manually searched for corresponding publications for each repository and excluded all studies that lacked corresponding methods or any information to match sputum samples with sample donor information. BioProjects that were not demultiplexed before SRA submission were excluded from this study.

In total, 36 BioProjects passed our inclusion criteria. For each BioProject, we used the same pipeline. All function parameters were set to defaults unless otherwise noted. First, we pulled all associated sample fastq files. As studies sequenced different 16S regions using different primers, we standardize all sequences by applying a uniform quality filter (maxEE=5) and trimming the first 20bp and truncating to 140bp (trimLeft=20; truncLen=140) using the dada2::filterAndTrim function in R, with the remaining parameters set to defaults. Samples for which fewer than 20% of the reads passed this sequencing quality filter were discarded.

To optimize the dada2 error inference step, we included all samples that passed our filters, including non-sputum samples. We used dada2::dada to infer sequencing error rates and removed chimeras using dada2::removeBimeraDenovo. We assigned taxonomy by BLASTing each sequence against the NCBI 16S ribosomal RNA database, identifying the top 10 alignments, and reporting the most frequent genus call. All subsequent data handling was built using the phyloseq packages in R. We successfully analyzed 5201 samples using this pipeline and subsequently removed non-sputum samples such as sequencing controls and paired samples from other body sites, as well as non-observational (experimentally manipulated) samples from further downstream analysis.

The resulting dataset contained 4171 sputum samples with manually curated and matched sample metadata. Sputum samples contained a median of 20061 reads (median 19497, range 1045-1264313). DNA sequences were assigned to 1090 distinct genera. To mitigate study-specific sequencing bias, we only analyze genera that were detected across the majority (>18) of studies. This yielded 74 genera across our dataset for downstream analysis. To account for the variability in total read output for each individual study, all samples were then subsampled to a sequencing depth of 2000 reads; 145 samples were below this read threshold and thus discarded, yielding 4026 sputum samples across 1184 subjects.

### Unsupervised clustering models

To identify pulmotypes, we applied three unsupervised clustering approaches: Dirichlet Multinomial Mixture (DMM), Partitioning Around Medioids (PAM), and K-means. For all models, only the earliest available sputum sample for each subject was included during model fitting to avoid individual over-representation.

### Dirichlet Multinomial Modeling

Following the procedure outlined in Holmes et al. (Holmes et al., 2012), we partition samples into pulmotypes using Dirichlet Multinomial Mixture (DMM) modeling. DMMs have been used to perform unsupervised clustering on microbiome datasets across numerous contexts(Costea et al., 2018, Holmes et al., 2012, Widder et al., 2022). We fit DMMs using the DirichletMultinomial R package. Cluster assignment confidence was calculated as the likelihood of observing a given sample in its maximally likely component in the fit DMM model.

### Partitioning Around Medoids (PAM)

We implemented Partitioning Around Medoids (PAM) clustering using the *cluster* package in R, based on the approach introduced by (Kaufman and Rousseeuw, 2009). PAM has demonstrated utility in microbiome studies across multiple settings (Arumugam et al., 2011, Sun et al., 2025, Acosta et al., 2018). Optimal cluster number was selected by maximizing the Calinski–Harabasz (CH) index.

### K-means model

K-means clustering was applied as described by (MacQueen, 1967) through the base R ‘stats’ package, similar to other microbiome studies (Hampton et al., 2021, Boutin et al., 2017). We systematically tested different values of *k* (e.g., 2 to 20) and applied the “elbow method” to determine the optimal number of clusters (the “elbow” in the plot of within-cluster sum of squares versus *k*).

### Average Silhouette Width (ASW)

Following the procedure introduced by Rousseeuw(Rousseeuw, 1987), we measure cluster separation by computing the **Average Silhouette Width (ASW)**. ASW captures how similar each sample is to its own cluster relative to the nearest alternative cluster, and then averages this value across all samples.

### Compositional differences

We test for compositionally distinct studies using a leave-one-out approach. Analysis of similiarty (ANOSIM) tests were performed on Bray-Curtis distances between each study and the composite of all other studies using the anosim function in the vegan R package(Oksanen et al., 2023). We then aggregate studies by reporting clinical center and perform both leave-one-out and pairwise center comparisons using ANOSIM. We also test pulmotype compositional differences using pairwise ANOSIM tests. For each set of comparisons, significance was assessed at p=0.05 (Bonferroni-corrected). Meaningful effect sizes were assessed at R>0.4 (Roberts et al., 2008).

## Supplementary Materials

**Supplementary Table S1.**
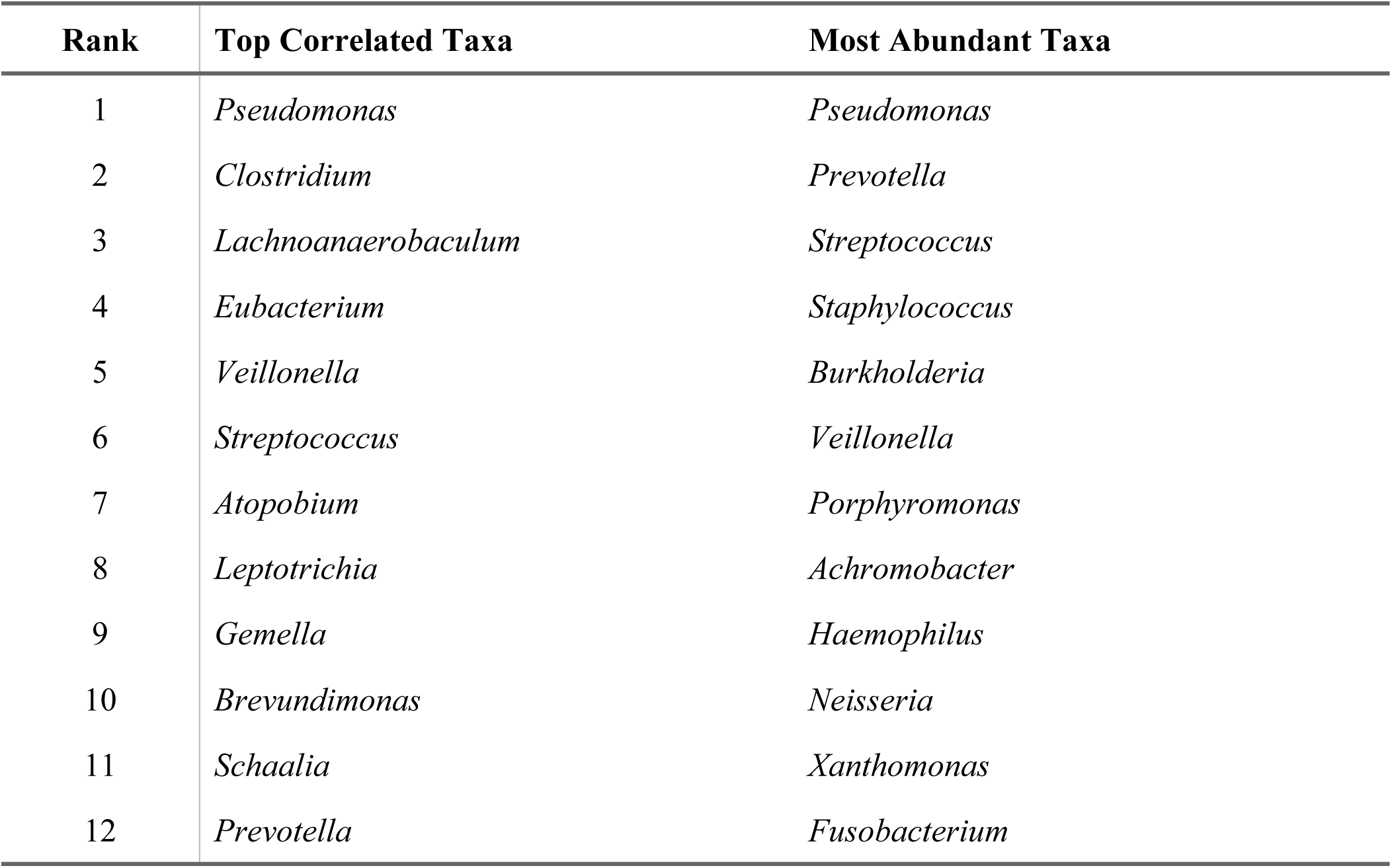
Taxa included in taxon-based models for predicting ppFEV₁ Taxa are ranked by their correlation with ppFEV₁ (Top Correlated Taxa) or by their mean relative abundance (Most Abundant Taxa) in the dataset. The taxon-based prediction models (Figure 9B) use the top 1, 2, 5, 8 or 12 taxa by rank order from the lists below.

| Rank | Top Correlated Taxa | Most Abundant Taxa |
| --- | --- | --- |
| 1 | *Pseudomonas* | *Pseudomonas* |
| 2 | *Clostridium* | *Prevotella* |
| 3 | *Lachnoanaerobaculum* | *Streptococcus* |
| 4 | *Eubacterium* | *Staphylococcus* |
| 5 | *Veillonella* | *Burkholderia* |
| 6 | *Streptococcus* | *Veillonella* |
| 7 | *Atopobium* | *Porphyromonas* |
| 8 | *Leptotrichia* | *Achromobacter* |
| 9 | *Gemella* | *Haemophilus* |
| 10 | *Brevundimonas* | *Neisseria* |
| 11 | *Schaalia* | *Xanthomonas* |
| 12 | *Prevotella* | *Fusobacterium* |

## Notes

### Competing Interest Statement

The authors have declared no competing interest.

### Funding Statement

This study was funded by the Cystic Fibrosis Foundation (awards BROWN23I0, BROWN21P0)and the CDC (contract 75D30120C09782).

### Author Declarations

The study used ONLY openly available human data that were originally located at: https://www.ncbi.nlm.nih.gov/bioproject. All bioproject numbers are provided in Table 1.

### Summary of Updates

Addition of Figure 9 (explanatory power of cluster-based versus taxon-based statistical models of lung function).

